# Progression and management of diabetes in Indian settings with universal access to health care: Protocol and plans for CHIPS cohort study

**DOI:** 10.1101/2023.01.24.23284975

**Authors:** Anjali Kulkarni, TR Dilip, Yogesh K Shejul, Prashant Bhandarkar, Puja Goswami, Palak Sharma, Priti Patil, KS James

## Abstract

There is a demand for more comprehensive studies related to diabetes management in Indian settings covering; incidence, multimorbidity and complications in diabetes patients, clinical progression, medication, and treatment-seeking patterns. CHIPS study aims to bridge this research gap through a systematic analysis of the medical records maintained under an employees contributory health services scheme (CHSS). The CHSS based in an urban metropolitan area has 89,204 beneficiaries. The hospital information management system (HIMS) has records of lab reports, clinical summaries, prescriptions, and drugs and other medical consumables, supplied for every interaction with CHSS. Firstly, a cohort of 835 patients newly diagnosed as diabetic in the year 2011-2012 was identified from the HIMS. Their 10-year (2011-2021) medical history after getting diagnosed as a diabetic patient was elicited from the HIMS in a retrospective manner. For comparison needs another cohort of 1670 age-sex matched non-diabetic beneficiaries was created and similar 10-year medical history was created. A total of 144,511 lab records and 247,473 drug records from the HIMS for the period 2010-2012 were scrutinized to identify newly diabetic patients and their non-diabetic counterparts. The reconstructed 10-year medical history of these two groups will be used to investigate the burden of diabetes in the community, transitions from a non-diabetic and pre-diabetic to a diabetic, excess morbidity in diabetic patients, seasonal variation in glycaemic levels, association between glycaemic control and frequency of health care utilization, and COVID-19-induced temporal changes in glycaemic control.

## Introduction

Global trends and projections on the prevalence of diabetes indicate that it is one of the most hostile chronic health conditions threatening the quality of healthy life globally. The situation is similar in India, where 14 percent of women and 16 percent of men aged 15 years and above either have high blood sugar levels (>140 mg/dl) or take medicine to control diabetes. (1) According to the International Diabetes Federation, of the global estimate of 536 million people with diabetes in the 20-79 age group, 14 percent reside in India. (2) The number of persons with diabetes in India is estimated to be 93 million by 2030. Such an elevation in the number of diabetes patients would lead to an exponential increase in financial resources required to prevent, diagnose, and treat this non-reversible condition. Estimates show that diabetes accounts for 3 percent of deaths in India and that years of life lost (YLL) due to diabetes-induced premature mortality is more than years of life lived with disability (YLD) due to diabetes. (3). With the ongoing epidemiological transition these YLD will be more than YLL in future years, which underlines the need for more research and interventions for countering the comorbidities in the life cycle of patients diagnosed with diabetes.

As noted in most of the less developed countries, the prevalence of self-reported diabetes in India is relatively higher in the urban population than in the rural population and in higher socio-economic strata than in lower socio-economic strata (4–6). Though there are several studies on the self-reported prevalence of diabetes in India, only a few studies provide estimates of the incidence of diabetes in Indian settings. (7–9) These studies based on longitudinal surveys of population cohorts from various sites have illustrated the effect of various behavioural risk factors on the incidence of diabetes. Additionally, Indians are characterized by a stronger genetic predisposition to diabetes and a greater degree of insulin resistance. (5,9,10).

Diabetes is often accompanied by connected chronic comorbidities. Conditions like hypertension, hypertriglyceridemia, retinopathy, thyroid disorder, hypercholesterolemia, dyslipidaemia, cardiovascular disease, stroke, and chronic kidney disease, are prevalent in Indian diabetes patients. (10–16) Though comorbidities are part of diabetes, little is known about their onset in diabetic patients, which is essential for a better understanding of risk stratification for the incidence of these morbidities. Another significant issue is the lack of preparedness of the health systems to manage diabetes effectively. All these strengthen the call to fill the research gaps in understanding the incidence, disease burden, spectrum of complications, and management of diabetes in the country. (4,17)

This paper is a brief description of the protocol drafted for a scientific investigation into the onset and multiple aspects of the management of diabetes in the life cycle of patients in an urban community in Greater Mumbai. All the members of the community are beneficiaries of the contributory health services scheme (CHSS) provided by their government employer. The CHSS is collaborating with the International Institute for Population Sciences (IIPS) for this research study. The CHSS-IIPS project titled CHIPS study aims to generate scientific evidence that can contribute towards the strengthening diabetes management in India.

### Contributory health service scheme

The CHSS had 89,204 beneficiaries in the year 2021, which includes current employees, retired employees, and their dependent family members. Out of these, 49,030 beneficiaries resides in the well-planned gated township, and 40,174 beneficiaries living outside the township are spread over various localities in Greater Mumbai. CHSS offers medical care to the employee, spouse, and their children aged upto 25 years. Employees and their spouses are entitled for CHSS benefits even after retirement from the service till death or unless they opt out for some reason like migration, employee’s resignation etc. Employee’s dependent parents are also enrolled as beneficiaries of this CHSS. The CHSS covers all preventive and curative care needs of all beneficiaries.

Each CHSS beneficiary based on their residential pin code is attached to a CHSS zonal dispensary. Beneficiaries can walk-in to that facility where the family physicians attended to their health care needs. The beneficiaries also can avail services under specialist doctors’ advice whenever needed at a dedicated multispecialty hospital which is situated inside the township. Hospital and all dispensaries are connected with a computerised hospital information management system (HIMS). There are 16 such dispensaries and one multispecialty hospital with 390 beds. The hospital and dispensaries are manned by about 65 specialist doctors, 200 family physicians, and an adequate number of nursing and technical staff.

## Methods

### Cohort Design

A 10-year retrospective cohort data set is constructed using the medical records maintained for these CHSS beneficiaries. The uniqueness of CHIPS cohort is the universal access to health care in this urban community. Results from most of the cohort-level studies available are likely to have a bias arising out of extreme inequities in access to health care services in the country. (18–20) Factors like limited access to medical facilities, perception of disease, and treatment continuity (21–23) could have added to delay in diagnosis of diabetes leading to aggravation of diabetes-related complications. Universal access reduces the risk of undiagnosed diabetes in the study community and complications arising due to delay or discontinuity in treatment. A retrospective type of design is preferred due to irreversible nature of diabetes and to make use of existing medical records data already available for the study population. This cost-effective design has the potential to answer a plethora of patient and medical care provider level questions relating to the management of diabetes.

The proposed phase I design was prepared after taking stock of the secondary data from existing medical records maintained for the newly diagnosed diabetes patients availing the medical services. Further systematic prospective follow-up of the same cohorts on intrinsic issues related to the management of diabetes will be taken up in phase II, planned for the future. The specific objectives of phase I planned to be completed by December 2023 are to:

➢ Estimate the incidence of diabetes and disease burden in the study population using hospital medical records.
➢ To understand the probability of and the time duration for transitioning from a pre-diabetic to a diabetic.
➢ To study the clinical progression and major clinical events in the course of the disease.
➢ To determine the diabetes-related excess morbidity in the population.
➢ To validate the existing estimates of disability-adjusted life years (DALYs) due to diabetes available for India and to contribute towards the preparation of actual disability weight for diabetes-related DALYS in Indian settings.
➢ To understand the seasonal pattern in glycaemic levels in diabetic patients.
➢ To study the interrelationship between frequency of interaction with the health care provider and glycaemic control. This analytical exercise will examine the frequency of utilization of health care services (in terms of periodic review, frequency of blood glucose monitoring and medicines) and its relationship with the management of blood sugar levels in the CHIPS cohort.
➢ To determine COVID-19-induced temporal variation in glycaemic control of diabetes patients.

### Health Information Management Systems Data

Near-uniform protocols are used for completing the medical records of every beneficiary who visits the medical facilities under this CHSS. All relevant investigations offered to the patients and laboratory reports are available in the HIMS. Details of medicines issued to the patients from an in-house pharmacy and the prescription is available in HIMS. We extracted details about their visits to casualty, dispensaries, outpatient departments, In-patient discharge notes, medical case progress notes, laboratory investigations, medical prescriptions, and death summaries. Details of patients referred to outside health facilities for specialized treatment or other reasons are also captured in the HIMS. The other vital parameters of patients like body weight, and blood pressure etc. expected to be recorded by physician in the case-sheet during outpatient care visit were noted to be incomplete in HIMS. The digitization of medical records in HIMS was started in 1996; however connectivity to peripheral zonal dispensaries were extended in phases till 2009. Hence complete data on all interactions between each beneficiary and health facilities are available from 2010.

Each beneficiary is issued an individual Medical Record Department (MRD) identity number at the time of inception into the CHSS. To follow the ethical code of conduct in this research every patient’s MRD number was masked using an encrypted version, as per a pre-defined algorithm. Anonymous patient records with encrypted identity numbers were provided to the CHIPS study team. All study objectives listed as part of phase I of CHIPS study described here will be based on information available from the medical records of beneficiaries. There will not be any kind of primary data collection or interaction with patients in the phase I part of the study.

### Identifying diabetes patients from medical records

We used an operational definition to identify diabetes patients from the medical records of all beneficiaries who utilized CHSS in 2011and 2012. Medical records also contained information on all beneficiaries who were tested for diabetes as well as those receiving medication for diabetes. The CHSS beneficiaries with either glucose levels above cut-off (FPG>=126 mg/ dL or PPG>=200 mg/dL or HbA1c >=6.5%) or who had taken diabetes medication in that calendar year or in-patients with classic symptoms of hyperglycaemia or hyperglycaemic crisis, were identified and categorized as diabetic. For example (Fig 1), among the 18,325 patients who had either undergone diabetes testing or taken diabetes medication in 2011, 8566 were in the diabetic patient category.

**Figure 1:**
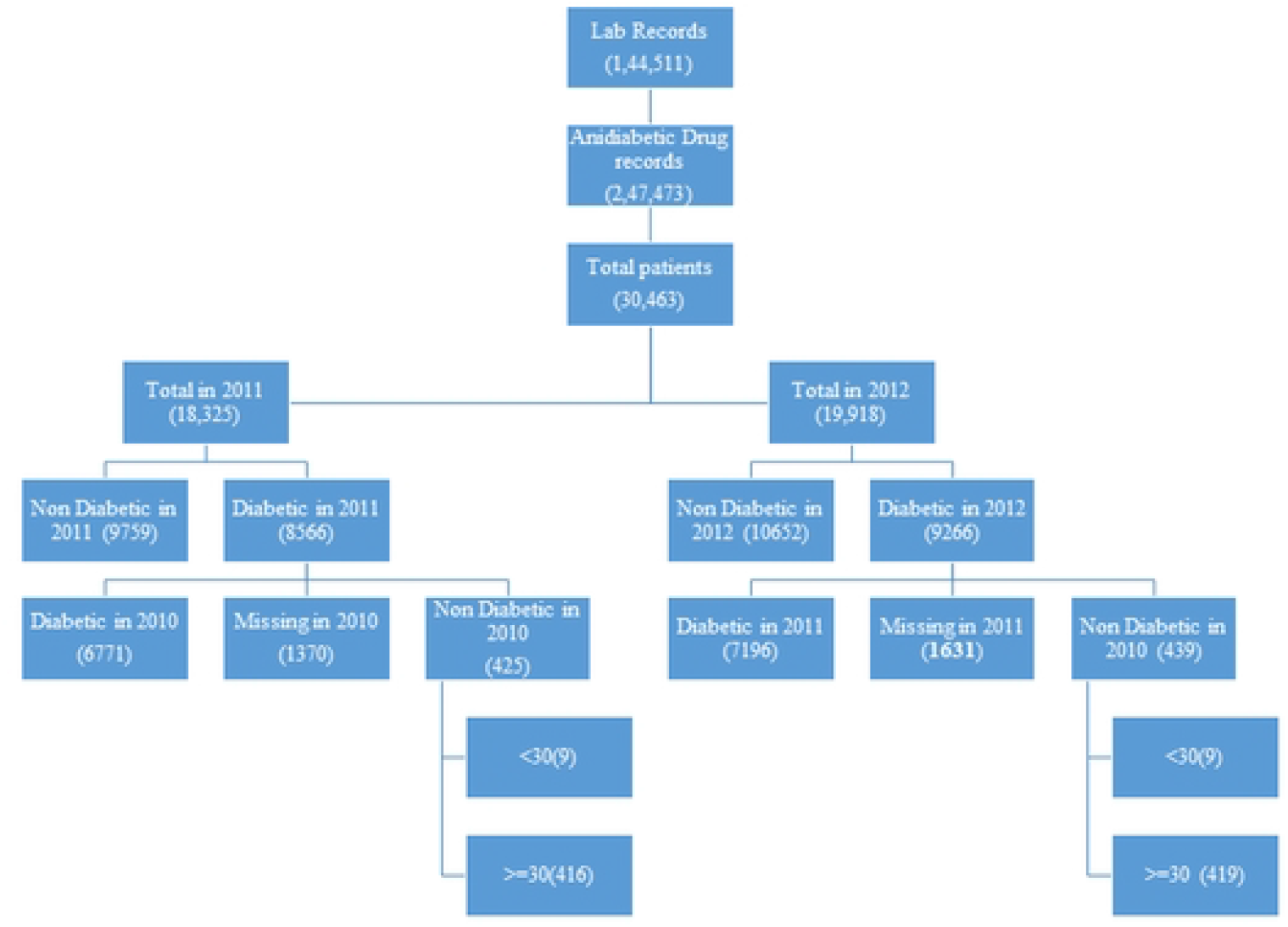
Flowchart on identification of 835 newly diagnosed diabetic cases from the medical records in 2011-2012

### Identification of newly diagnosed diabetes cohorts for follow-up

The next important step is the identification of newly diagnosed diabetes patient cohorts and eliciting their medical history from HIMS records through a retrospective follow-up in Phase I and prospective follow-up in Phase II planned for the future. Considering the availability of digitized data from HIMS we fixed the retrospective follow-up period for the newly identified patients to 10 years. Despite this data constraint, we consider a 10-year follow-up period is sufficient to study the diabetes-related comorbidity and excess mortality among the cohort patients. All those new diabetic patients diagnosed as diabetic in 2011-2012 were eligible to be in this cohort. Those who were found to be diabetic in a particular year but were tested as non-diabetic and were not taking any medicine for diabetes in the previous year, were defined as newly diagnosed diabetic patients. We scrutinized the entries in 144,511 lab records and 247,473 drug records available in HIMS for the period 2010 to 2012 to identify the newly diagnosed diabetic patients in 2011 and 2012. The figure 1 shows how 835 new diabetic patients (416 in 2011 and 419 in 2012) were identified for the 10-year retrospective follow-up.

### Identification of non-diabetic beneficiary cohorts for follow up

A separate cohort comprising of beneficiaries without diabetes at the time of initiation of original cohorts (i.e 2012) is required for comparison purpose for arriving at excess co-morbidity and for understanding transition from pre-diabetic stage in 2012 to diabetic in the 10 year follow-up period. For this an age-sex matched control group with double the size of newly diagnosed diabetes cohort was created from the same HIMS database. This group of 1670 non-diabetic CHSS beneficiaries were selected from list of persons who were tested as non-diabetic in 2012 and were not taking any medicine for diabetes.

### Cohort characteristics

In summary, the CHIPS cohort comprised of 835 newly diagnosed diabetic patients in 2011-2012, of which 55 percent were females (Table 1). The majority of males (83 percent) in CHSS were employees, while the majority of females (85 percent) were dependents of employees. This compostion is a reflection of the overall work participation among women in India. Age distribution of newly diabetic patients shows the onset of diabetes to be earlier in females than males. As can be expected the charecterastics of the age-sex matched control group of 1670 non-diabetic patients were similar to newly diagnosed diabetic cohorts.

**Table 1:** Baseline characteristics of CHIPS study subjects, 2011-12

|  | Newly diabetic patients (Cases)- % (N) |  |  | Non-diabetic beneficiaries (Controls) % (N) |  |  |
| --- | --- | --- | --- | --- | --- | --- |
|  | Females | Males | Total | Females | Males | Total |
| **Age in years** |  |  |  |  |  |  |
| 30-39 | 13.2 (61) | 5.3 (20) | 9.7 (81) | 13.2 (122) | 5.3 (40) | 9.7 (162) |
| 40-49 | 30.2 (139) | 18.2 (68) | 24.8 (207) | 30.2 (278) | 18.2 (136) | 24.8 (414) |
| 50-59 | 27.1 (125) | 30.5 (114) | 28.6 (239) | 27.1 (250) | 30.5 (228) | 28.6 (478) |
| 60-69 | 18.7 (86) | 25.7 (96) | 21.8 (182) | 18.7 (172) | 25.7 (192) | 21.8 (364) |
| 70-80 | 10.4 (48) | 17.9 (67) | 13.8 (115) | 10.4 (96) | 17.9 (134) | 13.8 (230) |
| >80 | 0.4 (2) | 2.4 (9) | 1.3 (11) | 0.4 (4) | 2.4 (18) | 1.3 (22) |
| **Type of beneficiaries** |  |  |  |  |  |  |
| Employees | 14.9 (69) | 83.2 (311) | 45.5 (380) | 16.9 (156) | 84.6 (633) | 47.2 (789) |
| Dependents | 85.1 (392) | 16.8 (63) | 54.5 (455) | 83.1 (766) | 15.4 (115) | 52.8 (881) |
| **Total** | **100 (461)** | **100 (374)** | **100 (835)** | 100 (922) | 100 (748) | 100 (1670) |

### Limitations

The charecteristics of a government employer based CHSS cohort will not be matching exactly with a population based cohort. Firstly the age composition in any similar CHSS cohort will be older than general population cohort. For this, the age-adjusted rates can be prepared for comparison of results from this study with other national or sub-national scenarios. Secondly, there will not be any representation for the very poor in this cohort as CHSS only comprises of salaried/ retired government employees and their dependents. Due to this factor, we expect the prevalence of diabetes to be relatively higher in this relatively more affuent CHSS population than in a general population.

### Attrition in the cohort size

The 10-year retrospective medical record data on the above-mentioned diabetes-related parameters were compiled for the 835 newly diagnosed patients. There were 586 patients who were available at the end of the follow-up period in 2021. The year-wise attrition in sample size or loss to follow-up cases in the cohort is presented in figure 2. Of the 249 patients lost during follow-up, we discovered that 78 patients had expired. The details of causes of death including whether it was diabetes-related death or otherwise will be obtained from their death records. Another 36 beneficiaries have left the CHSS and cannot be followed up. About 159 of the remaining patients were not seeking CHSS health services and the reason for the same is being traced as part of the research project. We expected mortality, gestational diabetes cases, migration of beneficiaries and discontinuation of health services as potential reasons for this attrition. The causes of attrition will be thoroughly investigated during the validation and processing of the medical records data.

**Figure 2:**
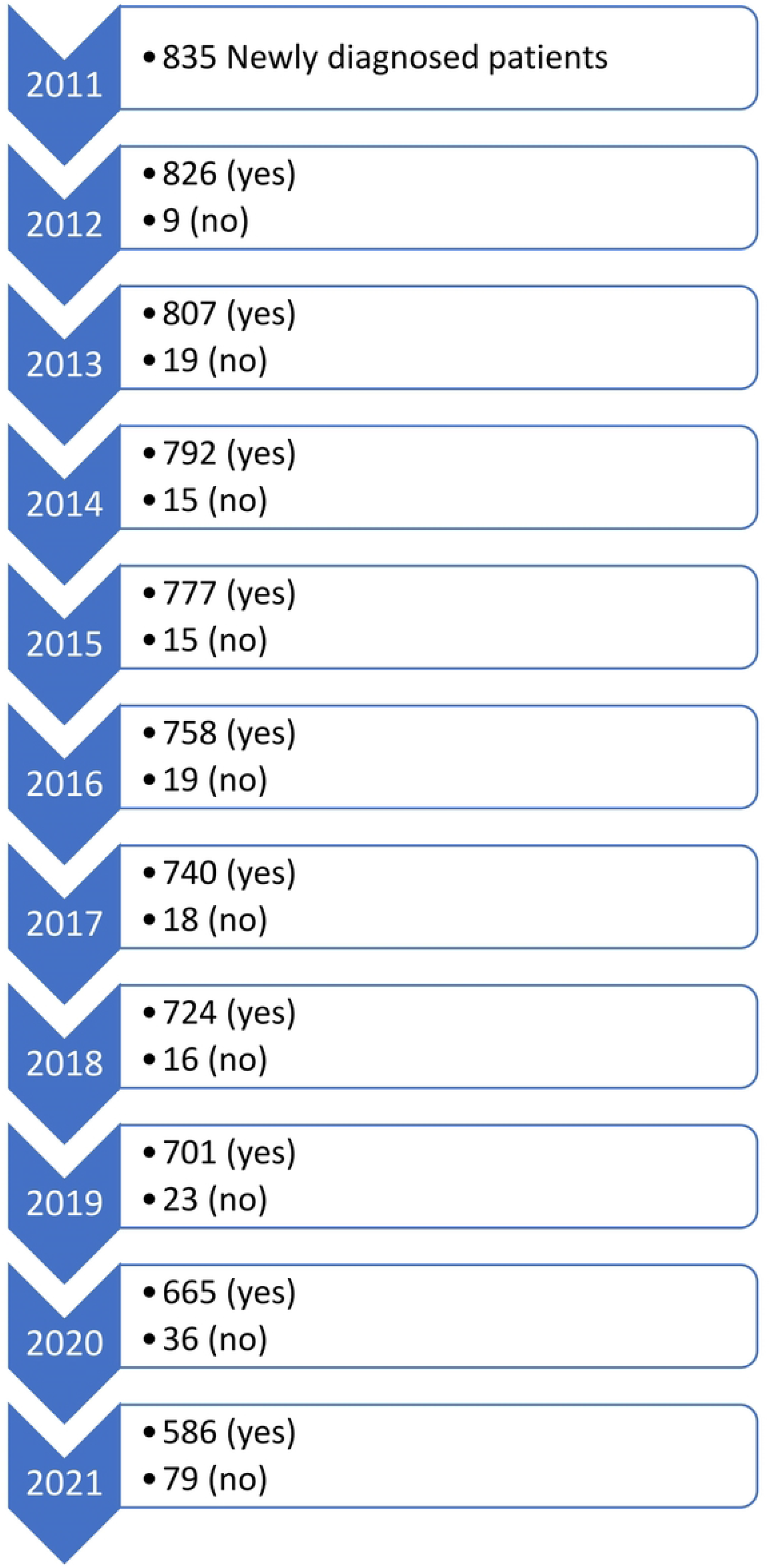
Attrition in the cohort of 835 newly diabetes patients followed up from 2011 to 2021

## Conclusion

The near-uniform protocols for patient evaluation and follow-up in the CHSS are the major strength of the CHIPS study. The results from this research study in a population with universal coverage will provide unbiased estimates of disease progression in the life cycle of the patients, including diabetes-related comorbidity, to the larger medical research community. This is important as quite often the economic and physical access to treatment alone doesn’t ensure better management of disease in the Indian context. Several cultural and gendered roles of patients within their family determines their capacity to seek treatment and pursue medical advice. Further, the database with complete details on medications and diagnostics prescribed and consumed by diabetic patients is a rich and unique resource for medical scientists working in the area of diabetes. We expect that the CHIPS study and its database has the potential to answer several policy-related research questions as well as capable of providing the supporting evidence for strengthening the guidelines for the management of diabetes in the country.

## Data Availability

All relevant data are within the manuscript and its Supporting Information files

## Acknowledgements

The authors are grateful to Dr Cherian Varghese, World Health Organization for his valuable comments and suggestions in the earlier draft of this paper.

## Ethics and dissemination

The Bhabha Atomic Research Centre (BARC) Hospital Medical Ethics Committee has given clearance for the Phase I of this project. Patients’ anonymity is assured in the CHIPS database by encrypting patient identification numbers in the HIMS database. Research findings will be disseminated among policy planners and stakeholders through a national workshop and through conferences and publications in scientific journals.

## Author contributions

AK, TRD, YKS and KS were involved in conceptualizing the design of the cohort. TRD, PG and PS performed the literature reviews. PB, PG, PS, PP, YKS and TRD were involved in compiling electronic medical records, cleaning the data and converting it into cohort-level data. TRD, PG, PS, YKS and AK authored the first draft of the paper. All other authors provided critical inputs for revising and finalizing the paper.

